# Variant Scientists in Genomic Medicine: Survey of Demographics, Training, Duties, and Professional Development

**DOI:** 10.1101/2025.01.28.25321277

**Authors:** Alexa Dickson, Kelsey R. Cone, Barbara K. Fortini, Jennifer Goldstein, Michelle L. Thompson, Matheus V.M.B. Wilke, Anna C.E. Hurst, Molly C. Schroeder, Katarzyna Polonis, Kevin M. Bowling

## Abstract

**Purpose:** Genomic testing has proven utility in disease diagnostics, guiding clinical management and improving outcomes. Use of high throughput sequencing by clinical laboratories has generated opportunities and challenges in data analysis resulting in the emergence of a laboratory role termed Variant Scientist. The aim of this study was to characterize this laboratory role.

**Methods:** We developed a thirty-item survey to collect information describing the current demographic landscape, salary ranges, work environments, training options, and professional development of Variant Scientists. The survey was disseminated to individuals conducting variant analysis in the United States from November 6, 2023, to March 15, 2024.

**Results:** Survey responders (n=87) are predominantly female (78%), age 40 or less (64%), hold advanced degrees (92%), and report four or more years of experience (75%). Responders report involvement in a diverse set of laboratory tasks and received relevant training on the job (78%). This workforce is satisfied with their career path (70%) and reports adequate support from employers. They note that resources and recognition from professional organizations are currently lacking.

**Conclusions:** Variant Scientists are a group of diverse genetics professionals that are highly educated and perform a variety of complex tasks within a clinical genomics laboratory.

## INTRODUCTION

Advances in genomic testing (particularly sequencing) along with decreasing costs has driven the rapid expansion of genetic testing in both clinical practice and research (1–4). With the widespread adoption of high-throughput sequencing, clinical laboratories now face significant challenges in tertiary data analysis due to the large volume of data generated. Tertiary analysis includes variant annotation, filtration, curation, and prioritization, typically in the context of the patient’s clinical presentation (5). Next, standardized classification of prioritized variants, following established guidelines and recommendations, must be conducted to provide biological context and determine clinical significance (6). This comprehensive analysis of potentially relevant genes and variants (which also includes case- and variant-level quality control) frequently requires labor-intensive expert evaluation of published literature and other available resources to ensure accurate clinical interpretation (5–7).

Historically, board-certified clinical laboratory geneticists (with the assistance of laboratory technicians) have performed data analysis and interpretation. However, the volume of generated sequence data has far outpaced the ability to interpret it within laboratory-established turnaround times without additional aid. This urgent need for analytical support has resulted in the creation of a specialized workforce often referred to as Variant Scientists (or Biocurators). Though Variant Scientists may be involved in a variety of different laboratory tasks, they are not certified to conduct clinical sign-out of patient reports or clinical assays, nor are they qualified to direct a clinical laboratory.

A review of available job openings for Variant Scientists (data not presented) suggests that clinical laboratories frequently seek candidates with at least a master’s degree level of education, ideally with considerable experience in variant classification. Additionally, expected or desired responsibilities for these roles may extend beyond genetic sequence analysis to include, but are not limited to, technical review of sequence data, support for improving existing workflows, and supporting the development and validation of new genomic assays.

Despite an apparent demand for Variant Scientists, little is currently known about this workforce. Therefore, we developed a survey to collect data from individuals who are currently working as Variant Scientists in the United States to assess the current professional landscape. Our study provides information about the demographic structure of this profession, levels of education and training, range of responsibilities, salaries, work environments, and job and career satisfaction. Our goal is to characterize Variant Scientists (e.g., who they are, what they do, and their contributions to genomic medicine) and provide data to support the development and sustainability of this workforce in clinical genomics. The results of this survey will be of interest to individuals currently working as Variant Scientists, individuals interested in a potential career in variant science, and to laboratory directors looking to Variant Scientists for assistance with maintaining efficient and growing operations.

## METHODS

### Survey development

The survey was developed by an internal working group at Washington University in St. Louis (Missouri, United States) consisting of clinical variant scientists, genetic counselors, and clinical laboratory geneticists. The 30-item survey consisted of multiple-choice, Likert scale, and short-answer questions to systematically collect information related to the role of the Variant Scientist (Supplemental Document 1). Survey questions encompassed basic demographics, education, training, professional experience, work environment, job title, salary, role diversity, and career satisfaction. The survey was hosted in REDCap and targeted individuals in the United States currently working as a Variant Scientist, or who serve in roles where variant analysis, curation, and interpretation is a significant part of their responsibilities. This survey was not intended for trainees or individuals holding, or eligible for, clinical laboratory or pathology board certification, such as laboratory genetics and genomics or molecular pathology.

### Survey dissemination

The survey was disseminated to 118 individuals via REDCap. The email distribution list was acquired via personal and professional networks. The survey was also distributed to members of the ClinGen Biocurators listserv (approximately 900 individuals). One reminder email was sent to each group of invitees. In addition, the ClinGen quarterly newsletter advertised the survey on December 20, 2023. The survey was opened for response between November 6, 2023, and March 15, 2024.

### IRB approval

Survey dissemination and anonymous data collection was approved by the Washington University in St. Louis Institutional Review Board (IRB ID: 202310106).

### Data analysis

Survey responses were summarized and analyzed using descriptive statistics (Supplemental Table 1).

## RESULTS

### Basic Demographics, Degrees, Certification, and Titles

In total, 87 survey responses were received. Survey response rate could not be calculated given methods of dissemination. Most survey responders were female (78%) and are 40 years or less in age (64%, Supplemental Table 1, Figure 1A). Most responders hold a master’s (38%, n=33) or doctoral degree (47%, n=41). Six individuals (7%) are medical doctors, with four of these individuals also holding a doctoral degree (Figure 1B). Seventy-six percent (n=66) of responders hold degrees in Genetics or Molecular Biology and 18% have been trained in genetic counseling (Figure 1C). Most responders hold no type of certification (62%). Of those who do hold certification, 22% are certified in genetic counseling and 11% report being certified by the American Society for Clinical Pathology. Most survey responders report holding positions titled Clinical Variant Scientist or Biocurator (66%), while the remaining responders identified as Genetic Counselors (8%), Staff Scientists (8%), Genome Scientists (6%), or hold a title related to a managerial or other role (12%; Supplemental Table 1).

**Figure 1.**
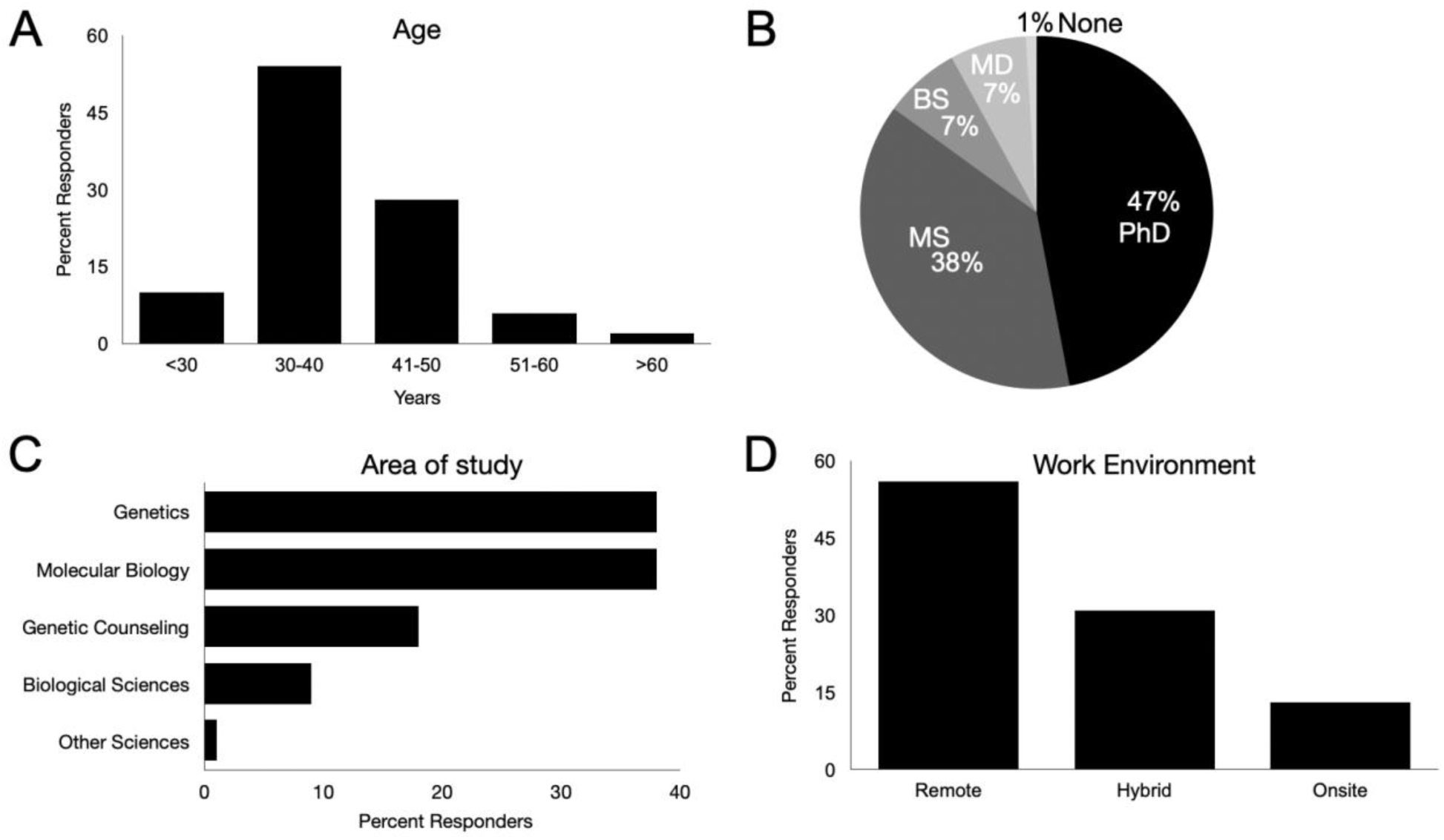
Demographics, education, field of study, and work environment of survey responders. A) Age of responders at time of survey. B) Terminal degree held by survey responders. C) Area of focused study. D) Reported work environment.

### Work Environment

Over half of survey responders report being employed by a commercial clinical laboratory (51%), with the remainder working in an academic clinical laboratory (33%) or translational research laboratory (16%). Only 13% of respondents report working fully onsite. In contrast, 56% indicate working remote and 31% report a hybrid work arrangement (Figure 1D).

### Years of Experience and Salary

Seventy-five percent of responders possess four or more years of experience in variant curation and analysis (Figure 2A; Supplemental Table 1). Notably, despite this moderate degree of experience, the majority have been in their current position for three years or less (58%). The most common base salary range reported by Variant Scientists is $76,000-$100,000 per year (33%). Variant Scientists with three years or less of experience earn less than $100,000 per year, while those with at least seven years of experience typically earn more than $100,000 annually (Figure 2B).

**Figure 2.**
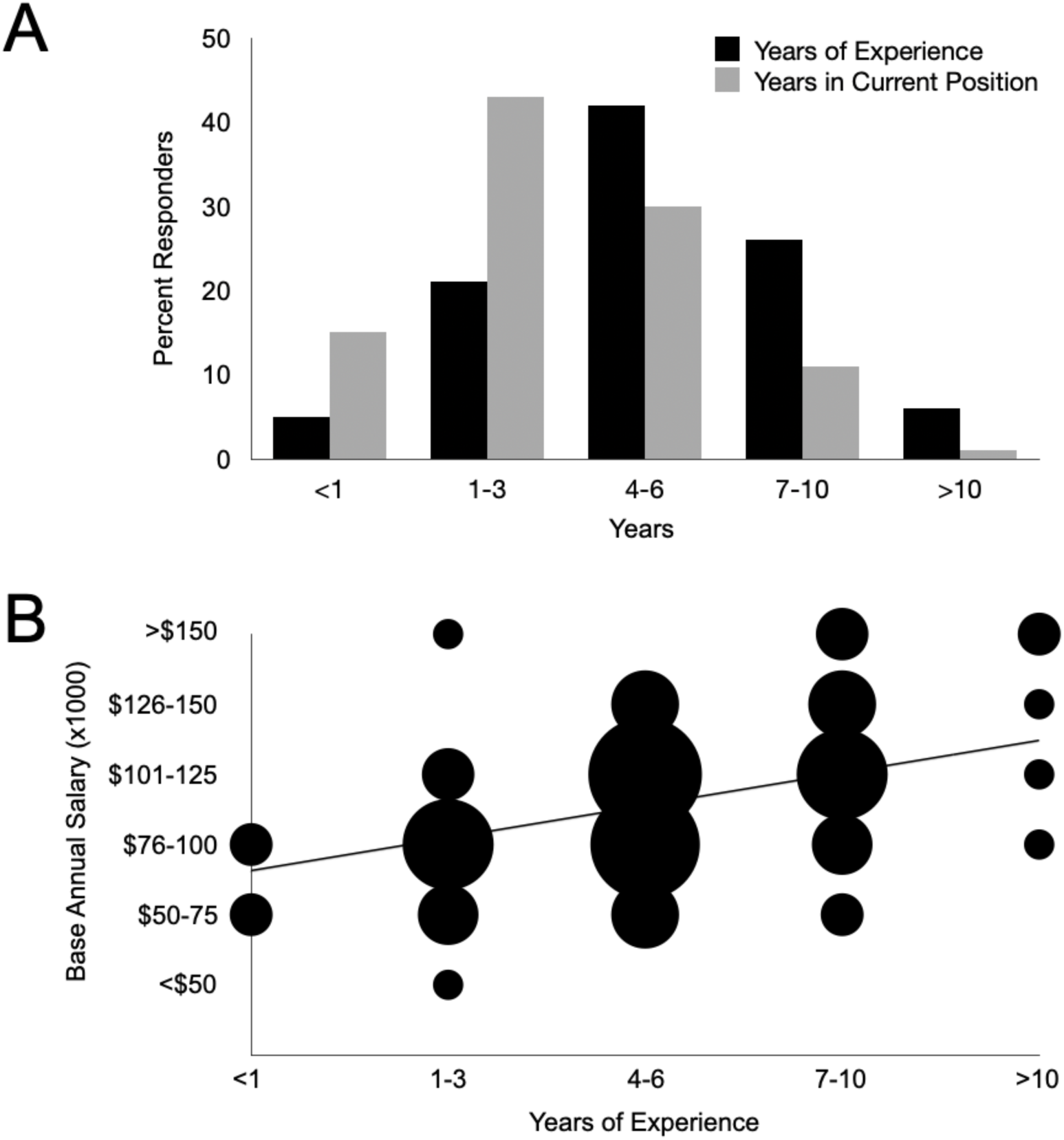
Professional experience, years in current position, and salary. A) 75% of survey responders possess four or more years of variant science experience while most (58%) have been in their current position less than three years. B) Variant Scientists with three years or less of experience typically earn less than $100,000 per year, while those with more experience (≥7 years) often earn more than $100,000 annually. Most Variant Scientists that completed the survey earn $76,000-$100,000 per year (33%). R^2^ = 0.169.

### Roles and Responsibilities

In addition to variant analysis, Variant Scientists often contribute to many facets of clinical laboratory operations, including test development and validation, quality assessment, management, and training. As part of the survey, we assessed Variant Scientist involvement in non-variant curation tasks (Figure 3). Notably, Variant Scientists are involved in training initiatives, with 85% reporting being often/sometimes involved with training others. This may include training novice variant scientists, junior analysts, or trainees completing laboratory fellowships. Variant Scientists also dedicate time to tasks related to data management/sharing (68% report involvement often/sometimes) which is essential considering the substantial data that is generated when conducting genomic testing. Moreover, many Variant Scientists report sometimes/often being involved with project management (56%) and managing/supervising others (46%). Variant Scientists report moderate involvement with clinical test development/validation and translational research efforts (Figure 3). Finally, the activities that survey responders report being least involved with include interactions with clients (i.e., ordering providers, 18%), genetic counseling (8%), and patient care (4%), as would be expected for those serving in laboratory roles primarily associated with variant analysis and interpretation.

**Figure 3.**
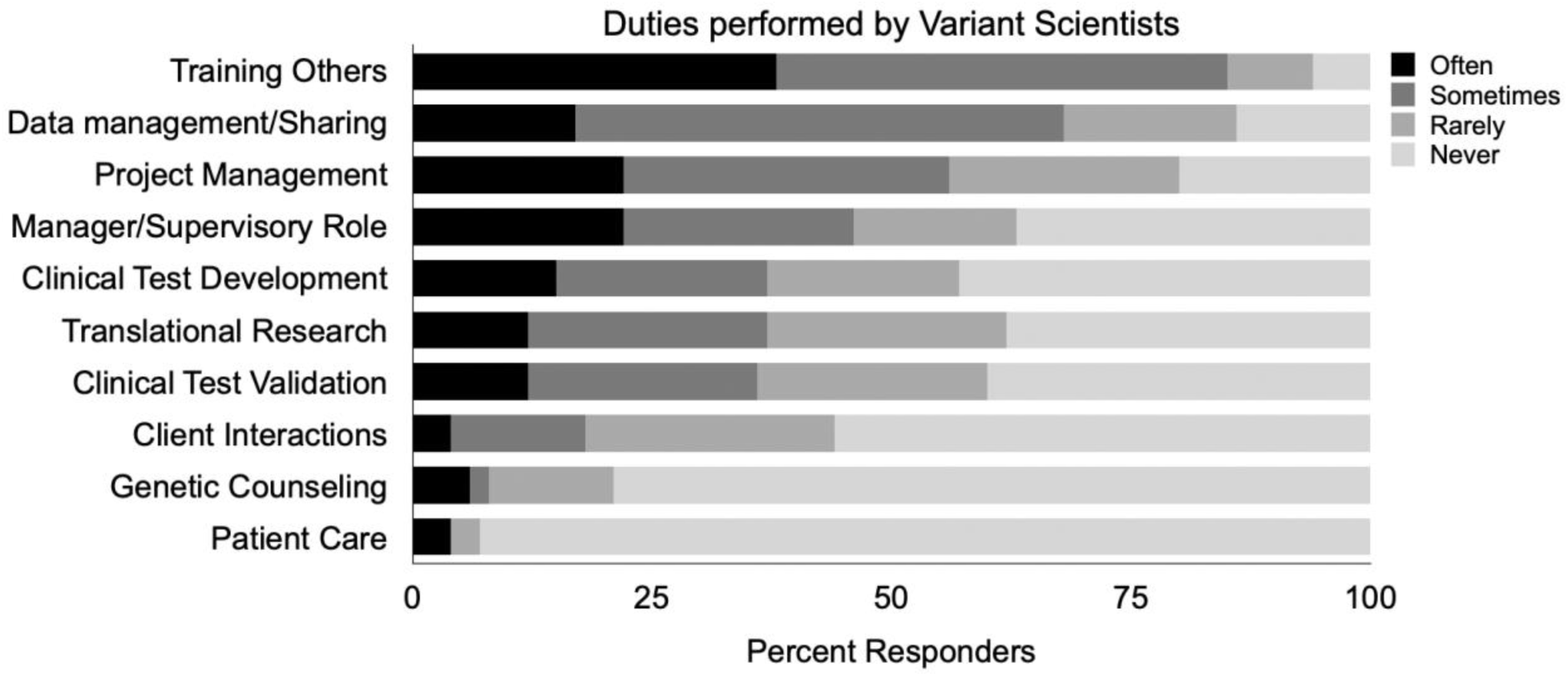
Assessment of the duties performed by Variant Scientists. Survey responders are most involved with training initiatives (85% of responders report often/sometimes involved) and data management/data sharing (68%). Survey responders report being least involved with (rarely or never) client interactions (i.e., ordering providers, 82%), genetic counseling (92%), and patient care (96%).

### Training and resources

Very few formal training programs currently exist that focus on development of individuals to conduct variant analysis, curation, and interpretation. Although, it should be noted that the College of American Pathologists has outlined requirements for individuals working in clinical laboratory settings including eligibility, competence, and continuing education. Four responders (5%) reported having completed a Variant Scientist training program. Of the 95% of responders that received no formal training, 78% (n=68) report on the job training, 11% (n=10) trained using Clinical Genome (ClinGen) resources, and 6% (n=5) report that they received relevant training as part of a postdoctoral fellowship (Figure 4). Most (87%) agree (strongly agree/agree) that they were adequately trained to perform variant analysis, curation, and interpretation.

**Figure 4.**
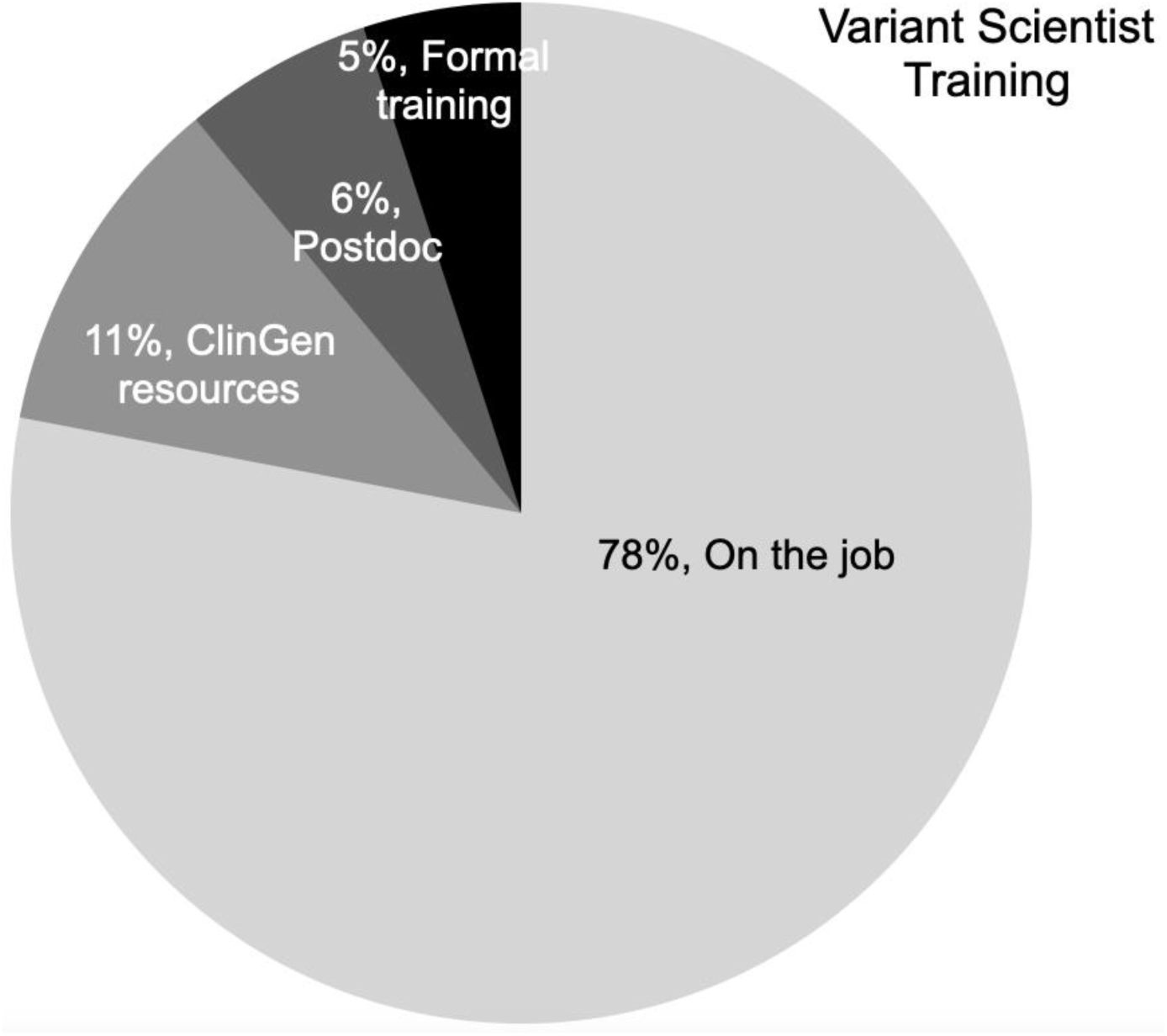
Variant Scientist training. Seventy-eight percent of survey responders report that they were trained on the job, 11% trained using ClinGen resources, 6% report that they received relevant training as part of a postdoctoral fellowship, and 5% report having attended a formal training program.

### Career Paths, Support, Professional Development, and Satisfaction

The career path of the Variant Scientist is varied and undefined as demonstrated by the diversity in reported job titles and daily tasks (Supplemental Table 1, Figure 3). Little standardization exists between those who employ variant analysts, likely due to this workforce representing a new and emerging career with limited visibility. However, survey data suggests that individuals working as Variant Scientists view their profession positively with most responders being moderately/extremely satisfied with their current position (86%), career path (70%), and career trajectory (61%; Supplemental Table 1).

In terms of resource allocation and support provided by their employer, 94% of survey responders indicate overall satisfaction with the technical resources (literature/database access, analytical tools, software, etc.) provided to effectively carryout position associated tasks (Figure 5). Seventy percent are also generally satisfied with employer-provided professional development resources, including provisions for continued learning opportunities and conference attendance. However, some responders report that mentorship, financial assistance for professional conference attendance, and continuing education opportunities are lacking within their specific institution/organization. In contrast, survey responders express a perception of relatively limited support from professional organizations with only 31% (strongly agree/agree) feeling that they provide relevant professional development resources to Variant Scientists. This perceived shortcoming includes facilitating continued learning initiatives and offering dedicated support for Variant Scientists. Only 11% of responders strongly agree/agree that professional organizations adequately recognize the role of Variant Scientist, highlighting the need for increased recognition, connection, and collaboration across the Variant Scientist community (Figure 5).

**Figure 5.**
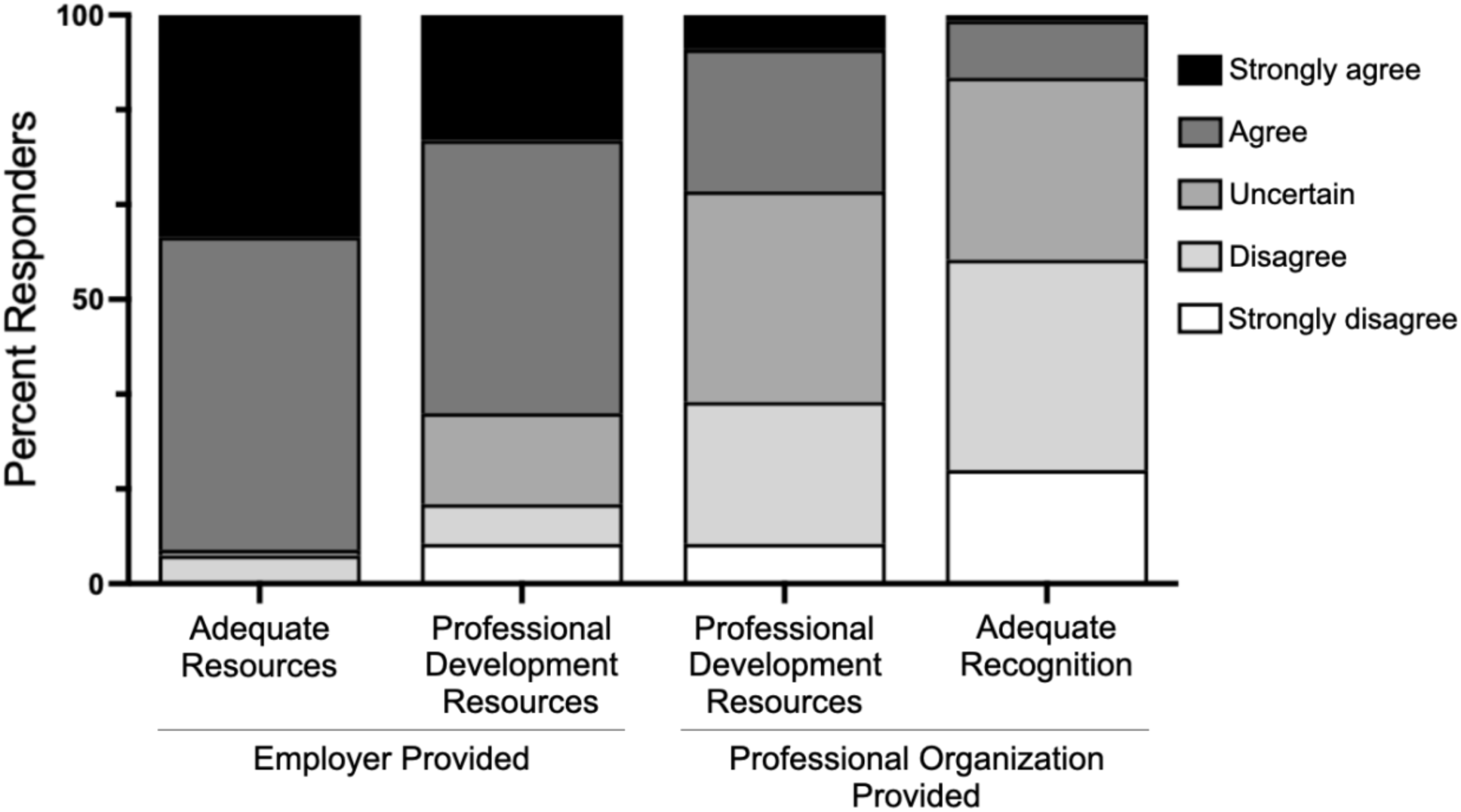
Variant Scientist support. Ninety-four percent of survey responders feel (strongly agree/agree) that their employer provides the technical resources need to perform their job, while 70% feel their employer provides relevant professional development resources. Thirty-one percent of survey responders feel that professional organizations provide relevant professional development resources to Variant Scientists, while only 11% of responders perceive that professional organizations adequately recognize the role of Variant Scientist.

## DISCUSSION

Most survey responders self-identified as female, consistent with other reports aimed at characterizing the medical genetics workforce (8–10). Responders report various professional titles, although the majority are in roles titled Clinical Variant Scientist or Biocurator. The diversity of reported job titles highlights the multifaceted nature of roles encompassing variant science, reflecting the interdisciplinary and collaborative nature of genetic research and clinical practice. Survey responders were also found to hold advanced degrees (master’s or doctoral degree) underscoring the specialized knowledge and skillset of the workforce. This result aligns with most present-day Variant Scientist job postings as they overwhelmingly require a doctoral degree in a relevant discipline or a master’s degree with at least some experience in variant analysis and interpretation.

In terms of work practices, our survey revealed that remote work is common in variant science as most responders report completely remote or hybrid roles. This observation points to increased remote/hybrid work practices within the field, reflecting the adaptability and flexibility inherent in contemporary professional settings and suggesting that laboratories are willing to support remote workers to obtain the needed analytical expertise.

Although Variant Scientists allocate significant portions of their time to variant classification, curation, and interpretation tasks (79% of survey responders agree/strongly agree this is the primary role of their position), their professional responsibilities extend beyond these core activities. This most often includes training others within the laboratory and tasks associated with data management and data sharing. Survey responders report being least involved with client (i.e., ordering provider) and patient interactions as is expected of those filling a primary laboratory role. Interestingly, only ∼35% of responders report being often/sometimes involved with new clinical test development and validation, suggesting that these tasks are typically driven by a clinically licensed laboratory director or other professional laboratory staff.

The results of our survey suggest that responders are moderately experienced, with 75% of responders reporting four or more years of experience, while 32% report seven or more years of experience. This finding is intriguing given that careers in variant science might be seen as a new occupational subspecialty and given the fact that so little was previously known about this workforce. Moreover, although survey responders typically report having worked in variant science for four or more years, 58% report having been in their current position for three or fewer years. This leads to questions about why individuals with variant science experience may have opted for a different position in the recent past. One plausible explanation is that professionals are seeking opportunities elsewhere in pursuit of professional and career growth, and/or increased salary. Alternatively, this observed trend could be influenced by recent fluctuations in workplace stability, including layoffs that have affected the clinical genetics community. Finally, in our survey results, experience level correlates with annual base salary and this correlation may be enhanced by individuals consistently seeking new opportunities, as aforementioned

Variant Scientists do not require, nor can they obtain, formal certification in variant science or interpretation. However, some individuals performing this role do hold certifications that are not Variant Scientist specific, such as American Society for Clinical Pathology certification. Moreover, 22% of survey responders report holding genetic counseling certification although only 8% of survey responders report actively working as a Genetic Counselor. This suggests that a considerable number of individuals trained in genetic counseling currently work as Variant Scientists, which is not surprising given that nine percent of genetic counselors report working in non-patient facing laboratory roles (11).

Given the technical and diverse nature of Variant Scientist positions, training is paramount to optimal and efficient job performance. However, only a small number of survey responders (5%) report having received formal variant/genome science training, while most survey responders report having been trained on the job (primarily learning through exposure and experience) or by ClinGen provided resources. Training of the variant science workforce through less formal means is to be expected given that currently only a few programs focus on the development of this specialized workforce. The training programs that do currently exist include short courses and employer-sponsored training modules for those already holding related genetics or biology degrees, curriculum embedded within related degree programs such as genetic counseling, or standalone master’s degrees in human genetics and genomics with a focus on variant science. In this varied landscape, there is no universally accepted curriculum standard or professional program accreditation available to compare or assess programs. This presents a major challenge for individuals that aim to pursue a career as a Variant Scientist (i.e., no well-defined path to entry) and for testing laboratories trying to identify and recruit individuals with the skills required for variant analysis. This lack of standardized training, competency, and continuing education puts the burden of establishment on individual laboratories which may lead to inefficiency and variable training outcomes. Regardless of training modality, lab-specific training will always be necessary as standard operating procedures, protocols, testing strategies, and reporting criteria may vary across laboratories. Nonetheless, more formalized training of the Variant Scientist workforce could reduce the current time investment required by employers to train new hires and would likely reduce the burden of onboarding.

As aforementioned, a considerable number of survey responders (11%) report having completed training using ClinGen resources. ClinGen, launched in 2013, represents a repository funded by the National Institutes of Health dedicated to building a central resource that defines the clinical relevance of genes and variants for use in precision medicine and research (12). ClinGen supports multiple different Variant Curation Expert Panels (VCEPs) composed of individuals with experience in a particular disease/gene area and biocurators who scour publications and public databases for relevant information (13). The majority of ClinGen’s curation work is carried out by volunteers who may benefit by gaining experience in variant classification, learning about a new disease area, interacting with experts in the field, and potential involvement with resulting publications. To support the growing number of volunteers, ClinGen has developed a series of variant curation training modules (14) to prepare individuals for involvement with a specific VCEP or that may be used by individuals that are either currently working as Variant Scientists or looking to enter the workforce. In addition to ClinGen, there are a variety of other existing resources and tools that may aid Variant Scientist training and day-to-day variant curation, analysis, and interpretation activities. We include in Supplemental Table 2 an extensive, but not exhaustive, list of resources that Variant Scientists provided when completing the survey. These resources are categorized based on their functionality and represent a diverse toolset that every clinical and translational genome scientist will find useful.

Individuals that completed the survey state that they are satisfied with their current position, career path, and career trajectory. Moreover, a majority report that they are satisfied by the support and resources afforded by their current employer, which is a positive indicator for laboratories that employ Variant Scientists, or at least those represented by the responders of this survey. In contrast, survey responders do not feel adequately supported or recognized by professional organizations. This highlights a need for resources facilitating career advancement (including potential pathways for certification and avenues for professional networking) and underscores the professional development aspirations among those working in variant science. Moreover, this demonstrates a perceived gap in the provision of resources and advocacy from professional bodies tailored to the specific needs of Variant Scientists, potentially warranting further attention and collaboration within the professional community. These factors should be carefully considered by relevant stakeholders, especially in the context of Variant Scientist recruitment, support, and retention.

While specific career ladders, promotional opportunities, and individual career progression for Variant Scientists were outside the scope of the survey, individual experiences and interaction with Variant Scientists provide a general, if limited, insight into these topics. Variant Scientist career progression seems to vary greatly, and the specific path is likely dependent on each individual and the opportunities afforded by their employer. These paths may be clinically focused (variant interpretation, report templating), technically focused (test development and validation, software and tool assessment), research focused (conducting analysis and testing for clinical trials or translational research projects), managerial focused (team management, supervisory roles, quality improvement initiatives), or represent any combination of these diverse paths. Any of these paths may lead individuals working in variant science to other career options (e.g., laboratory genetics and genomics fellowship) or they may choose to continue work as a Variant Scientist.

This survey presents several limitations that should be considered when interpreting results. The survey was developed by individuals who currently work, or previously worked, as Variant Scientists and this may introduce inherent bias in survey design. The survey was disseminated to individuals by personal networking (curated list of Variant Scientist email addresses) and via the ClinGen biocurators listserv; it is highly likely that many individuals doing relevant work may not have received the survey notification. Further, the survey was titled “Assessing the Variant Scientist occupational landscape” which may have resulted in it not being seen as relevant to individuals carrying out variant science roles (e.g., Genetic Counselors) but not holding the professional title of Variant Scientist. Finally, the survey was intended only for individuals working in the United States and should not be interpreted to represent the demographics or characteristics of the global or comprehensive Variant Scientist community.

We have for the first time characterized a cohort of Variant Scientists, which has not been done by any other group to date. While Variant Scientists primarily work to integrate information sources, curate, and interpret identified genetic variation in the context of the indication for testing, they also play crucial roles in training others, assessing quality metrics and analytical tools, managing workflows, curating resources, developing and validating clinical assays, and educating trainees. Interestingly, little has been known about the individuals fulfilling this role, and no professional organization currently represents this diverse, growing, and necessary workforce. It is our hope that the results of this survey increases the visibility of Variant Scientists and highlights their contributions to genomic medicine.

## SUPPLEMENTAL MATERIALS

Supplemental Document 1. IRB-approved version of the Variant Scientist survey that was disseminated to individuals currently working in variant curation, analysis, and interpretation.

## Data Availability

All data produced in the present study are available upon reasonable request to the authors.

## ACKNOWLEDGEMENTS

We are grateful to the Variant Scientists who completed the survey. We thank Yang Cao and Meagan Corliss for their thoughtful feedback throughout survey design, and Meredith Weaver and Maria DiStefano for help with survey dissemination via the ClinGen biocurators listserv. A special thanks to Kathy Dodds for assisting with the IRB protocol and REDCap design/distribution of the survey.

## ETHICS DECLARATION

The review board at the Washington University in St. Louis School of Medicine (IRB ID: 202310106) approved and monitored the study. Completion of the survey by participants was considered implied consent.

## AUTHOR CONTRIBUTIONS

Conceptualization: A.D., A.H., K.B.; Data curation: A.D., K.B.; Formal analysis: A.D., K.B.; Project administration: A.D., K.B.; Resources: M.S.; Visualization: A.D., K.B., M.T., M.W.; Roles/Writing - original draft: A.D., K.P., K.C., B.F., J.G., K.B; and Writing - review & editing: A.D., K.C., B.F., J.G., M.T., M.W., A.H., M.S., K.P., K.B.

---

**Supplemental Table 1:** Aggregate data representing select questions from the Variant Scientist survey.

|  |  | Number of Responders | Percent of Responders |
| --- | --- | --- | --- |
|  | Total Responders | 87 | --- |
| Gender | Female | 68 | 78% |
|  | Male | 19 | 22% |
| Age | <30 | 9 | 10% |
|  | 30-40 | 47 | 54% |
|  | 41-50 | 24 | 28% |
|  | 51-60 | 5 | 6% |
|  | >60 | 2 | 2% |
| Highest Degree Obtained | None | 1 | 1% |
|  | Bachelor's | 6 | 7% |
|  | Master's | 33 | 38% |
|  | PhD | 41 | 47% |
|  | MD | 6 | 7% |
| Field of Study | Genetics | 33 | 38% |
|  | Molecular Biology | 33 | 38% |
|  | Genetic Counseling | 16 | 18% |
|  | Biological Sciences | 8 | 9% |
|  | Other Sciences | 1 | 1% |
| Years of Experience | <1 year | 4 | 5% |
|  | 1-3 years | 18 | 20% |
|  | 4-6 years | 37 | 43% |
|  | 7-10 years | 23 | 26% |
|  | >10 years | 5 | 6% |
| Years in Current Position | <1 year | 13 | 15% |
|  | 1-3 years | 37 | 43% |
|  | 4-6 years | 26 | 30% |
|  | 7-10 years | 10 | 11% |
|  | >10 years | 1 | 1% |
| Work Location | Remote | 49 | 56% |
|  | Hybrid | 27 | 31% |
|  | Onsite | 11 | 13% |
| Laboratory Type | Commerical Lab | 44 | 51% |
|  | Academic Lab | 29 | 33% |
|  | Translational Research Lab | 14 | 16% |
| Certification | MB (ASCP) | 10 | 11% |
|  | CGC | 19 | 22% |
|  | Other | 4 | 5% |
|  | No Certification | 54 | 62% |
| Current Job Title | Clinical Variant Scientist | 48 | 55% |
|  | Biocurator | 10 | 11% |
|  | Genetic Counselor | 7 | 8% |
|  | Staff Scientist | 7 | 8% |
|  | Manager/Supervisor | 6 | 7% |
|  | Genome Scientist | 5 | 6% |
|  | Other | 4 | 5% |
| Base Salary per Year | Less than \$50,000 | 1 | 1% |
| | \$50,000 - \$75,000 | 13 | 15% |
| | \$76,000 - \$100,000 | 29 | 33% |
| | \$101,000 - \$125,000 | 27 | 31% |
| | \$126,000 - \$150,000 | 11 | 13% |
| | Greater than \$150,000 | 6 | 7% |
| Current Position | Extremely dissatisfied | 2 | 2% |
|  | Moderately dissatisfied | 4 | 5% |
|  | Neutral | 6 | 7% |
|  | Moderately satisfied | 39 | 45% |
|  | Extremely satisfied | 36 | 41% |
| Career Path | Extremely dissatisfied | 1 | 1% |
|  | Moderately dissatisfied | 12 | 14% |
|  | Neutral | 13 | 15% |
|  | Moderately satisfied | 40 | 46% |
|  | Extremely satisfied | 21 | 24% |
| Career Trajectory | Extremely dissatisfied | 1 | 1% |
|  | Moderately dissatisfied | 18 | 21% |
|  | Neutral | 15 | 17% |
|  | Moderately satisfied | 40 | 46% |
|  | Extremely satisfied | 13 | 15% |

**Supplemental Table 2:**
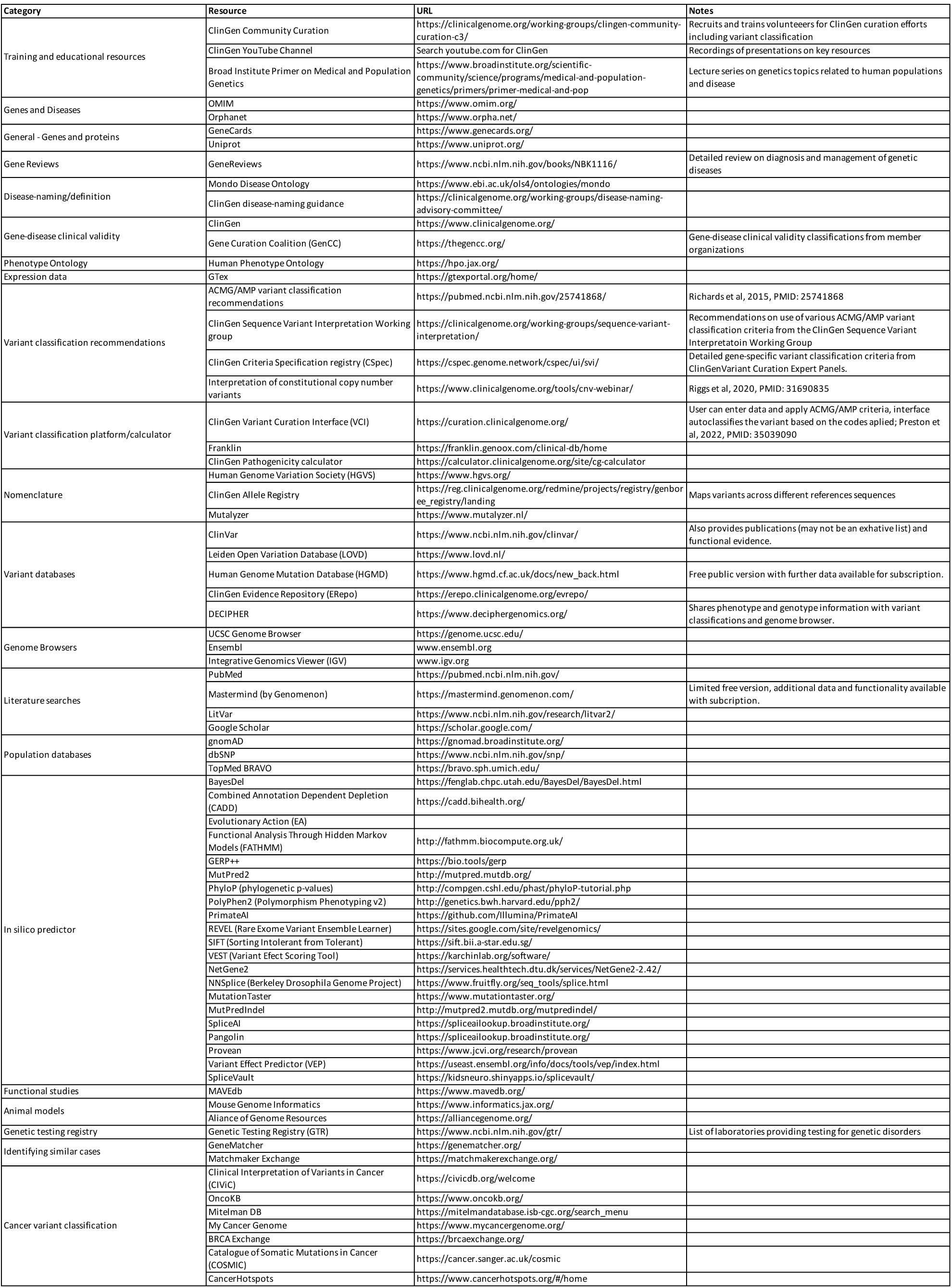
Resources for Variant Scientists. An extensive, but not exhaustive, list of tools and databases representing a diverse toolset that is relevant to Variant Scientist training and performance. The table is intended to provide a list of key resources that may be helpful in variant classification, but it is not exhaustive. All resources are freely available unless stated otherwise. Online searches for topics of interest may reveal additional resources. Please go to the websites listed for background information on each resource, available publications, and training.

## Assessing the Variant Scientist occupational landscape

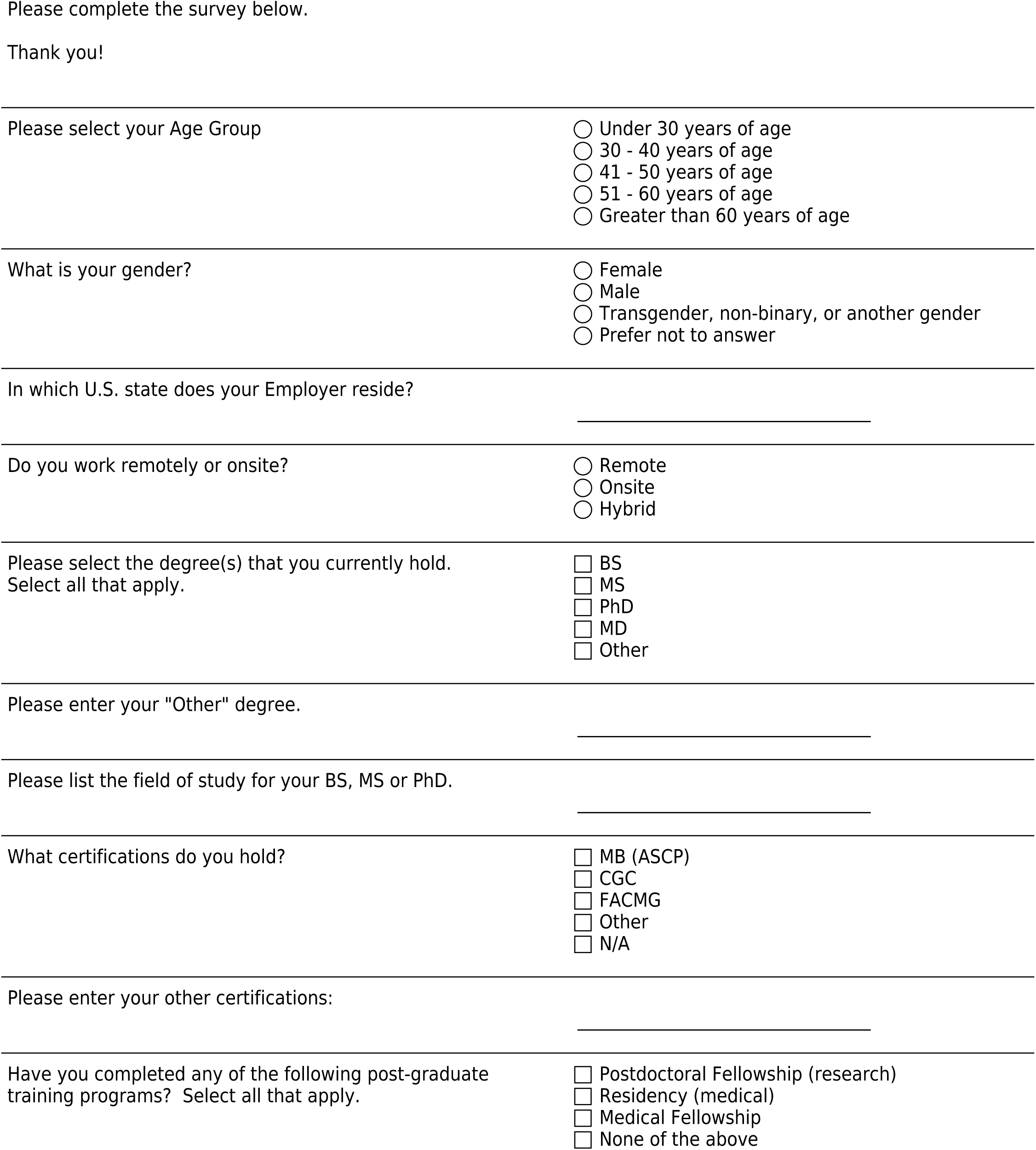
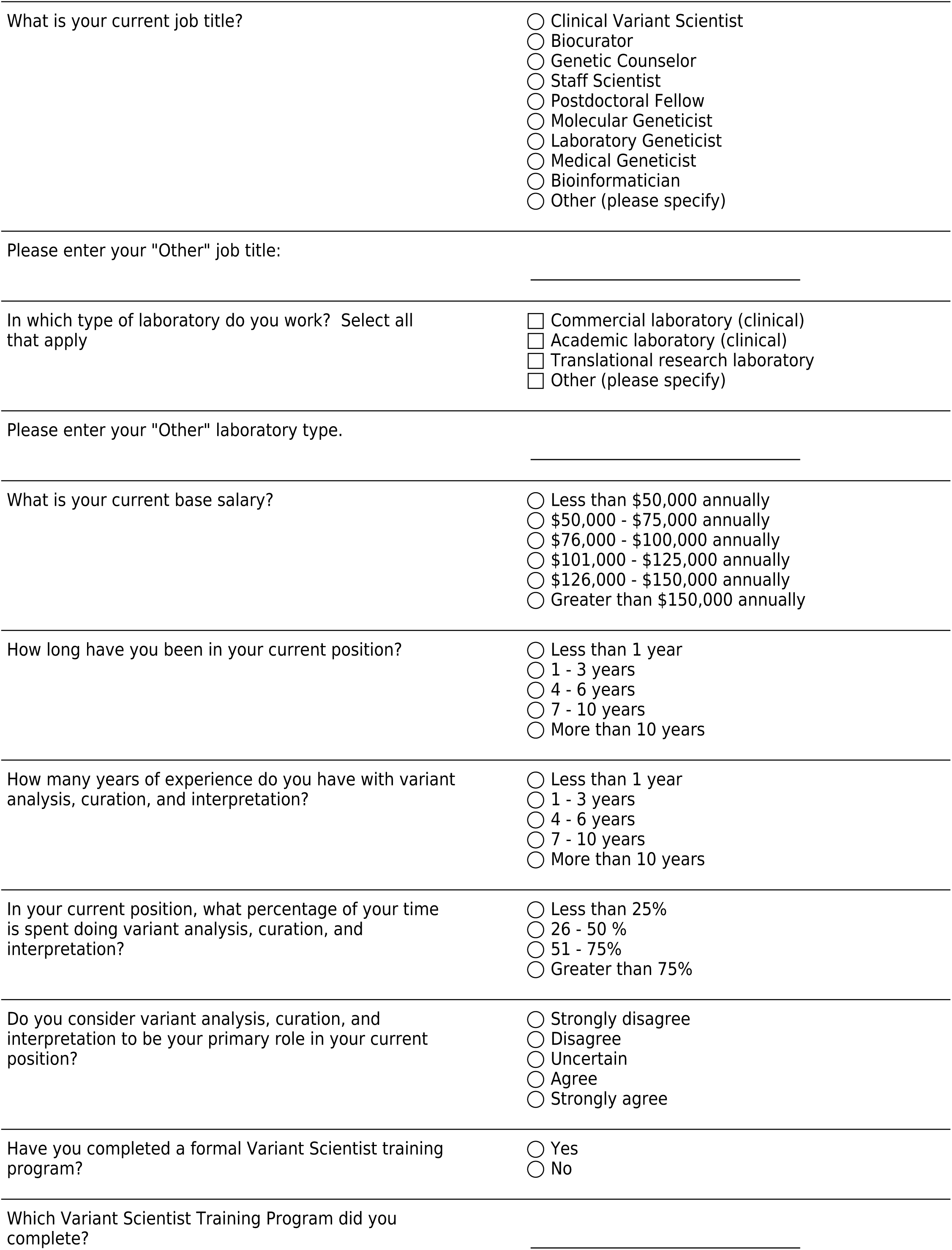
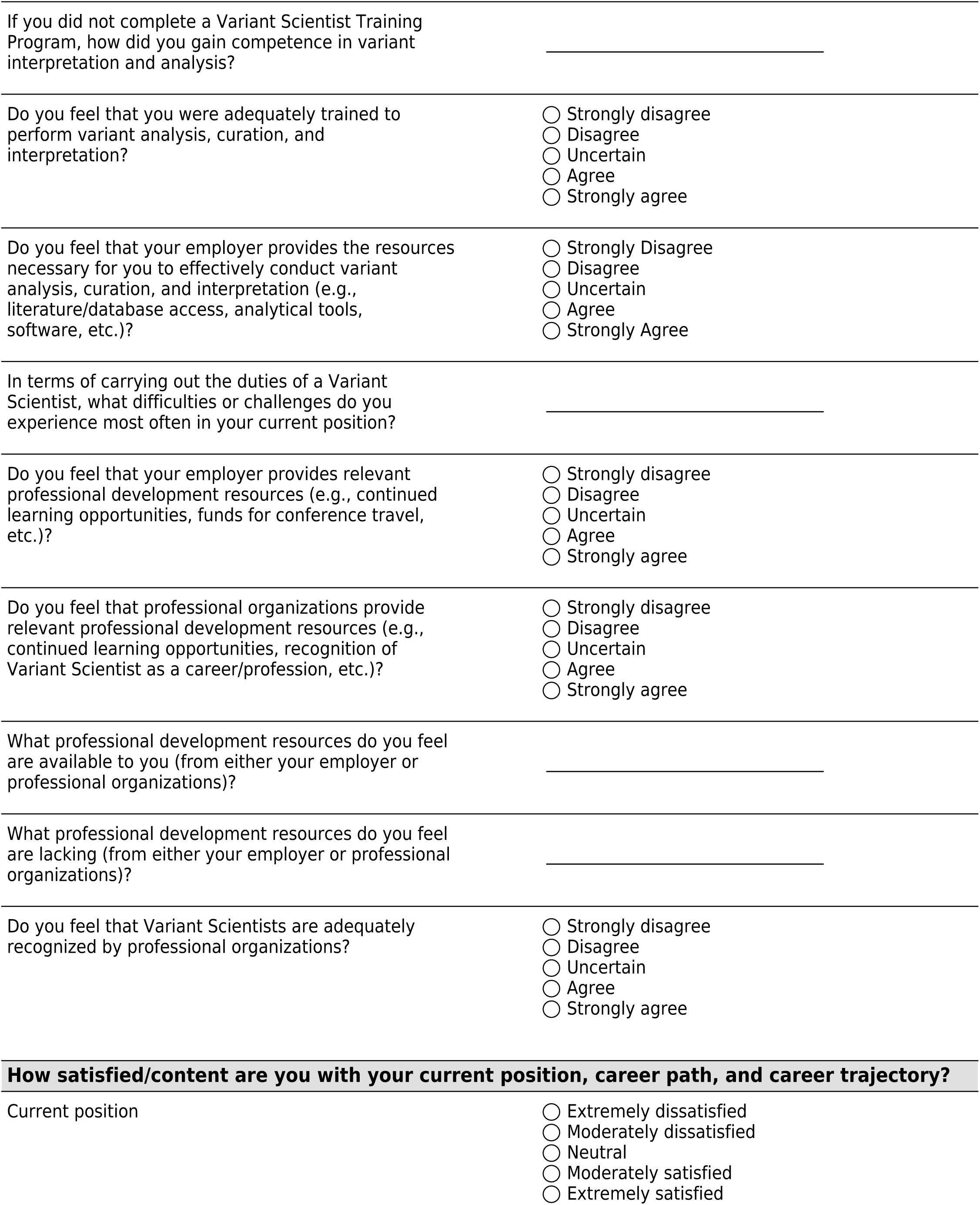
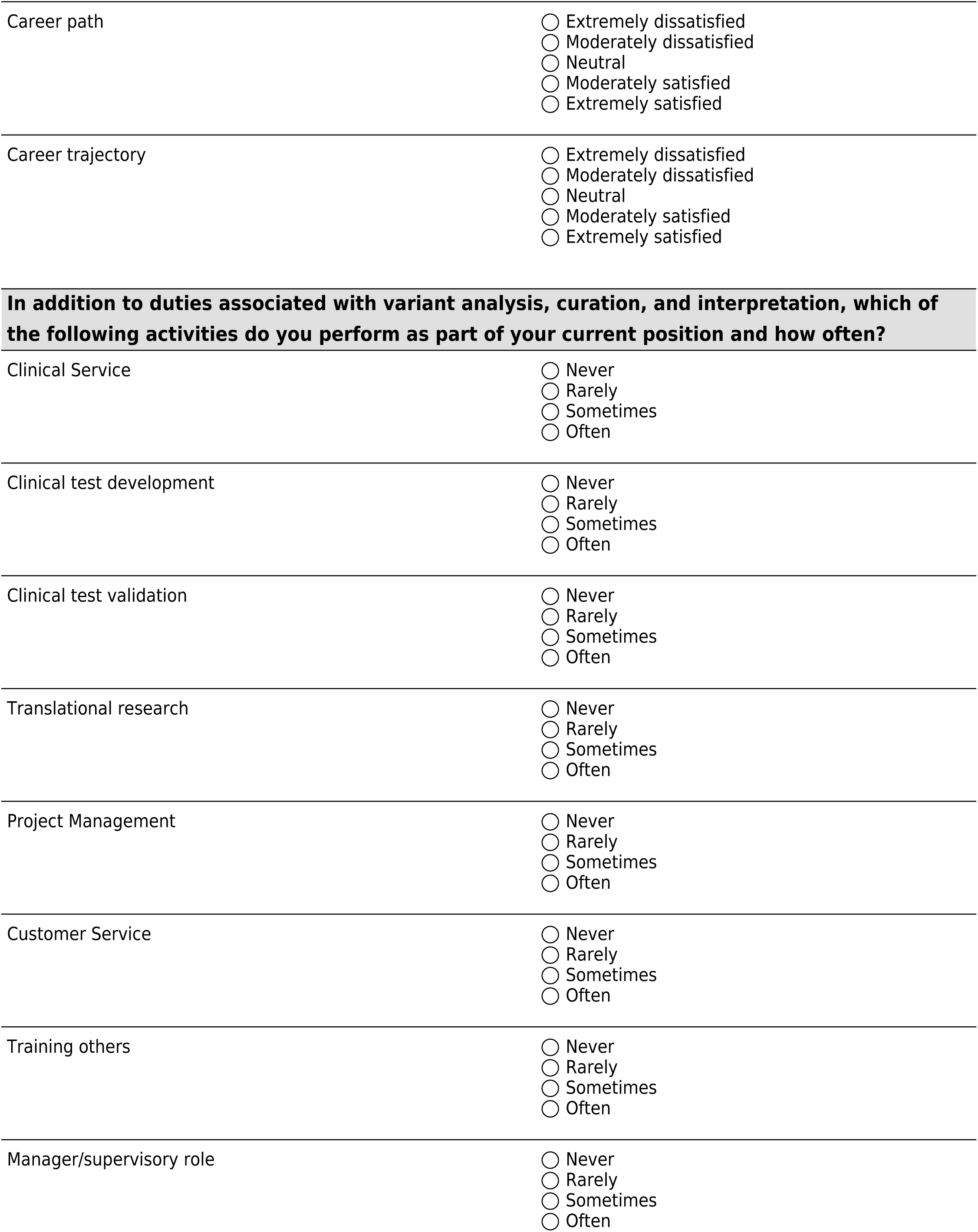
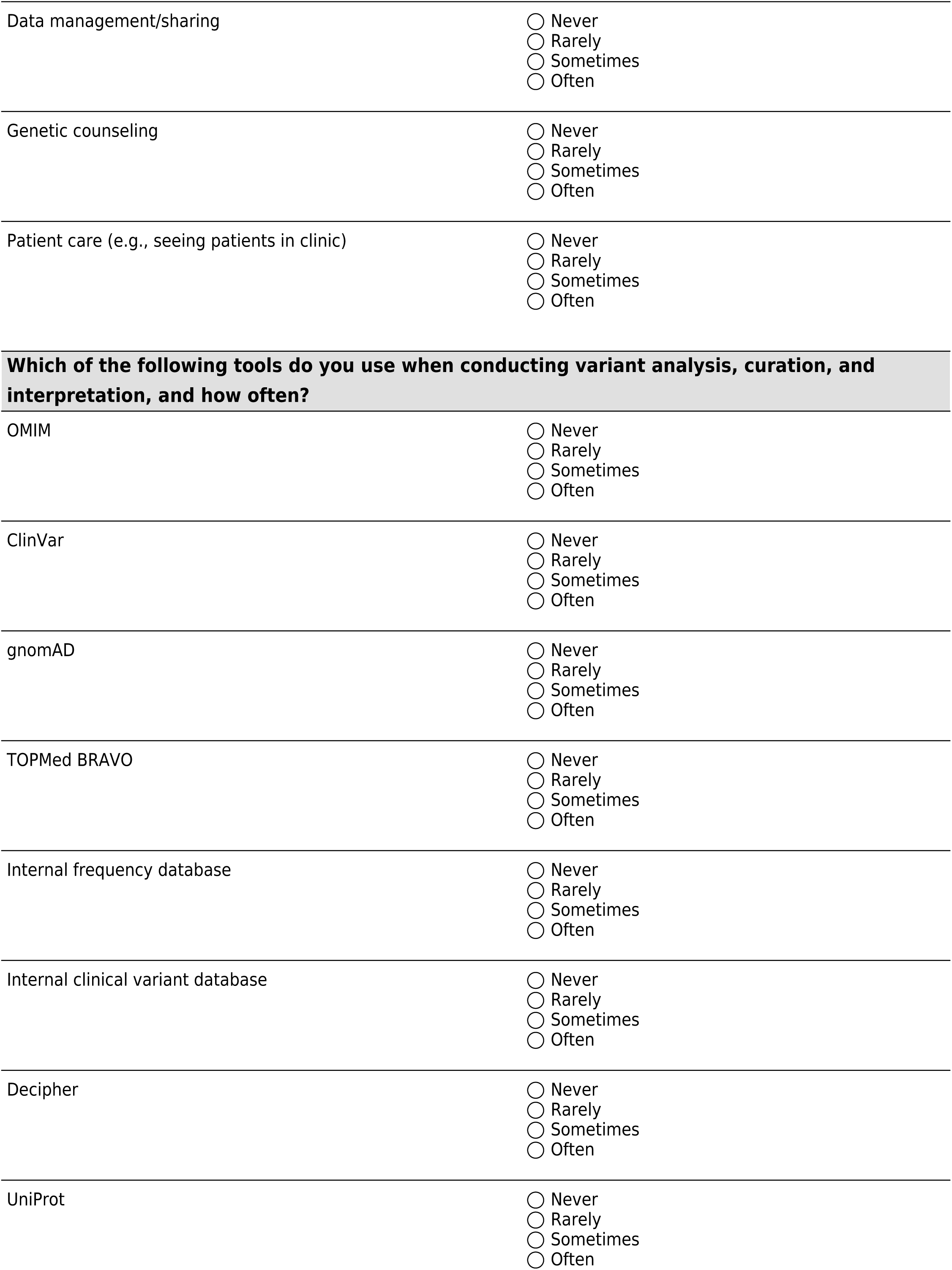
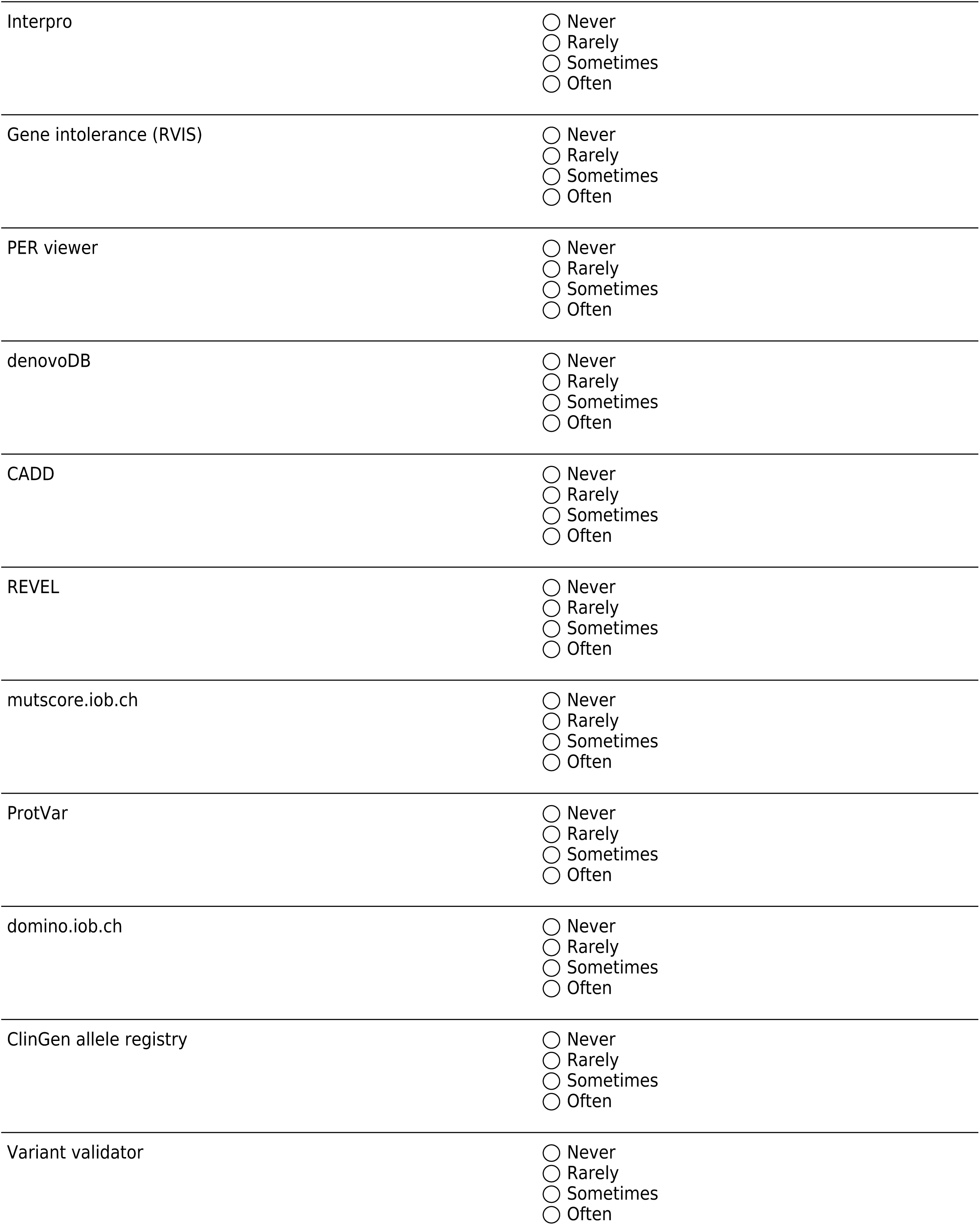
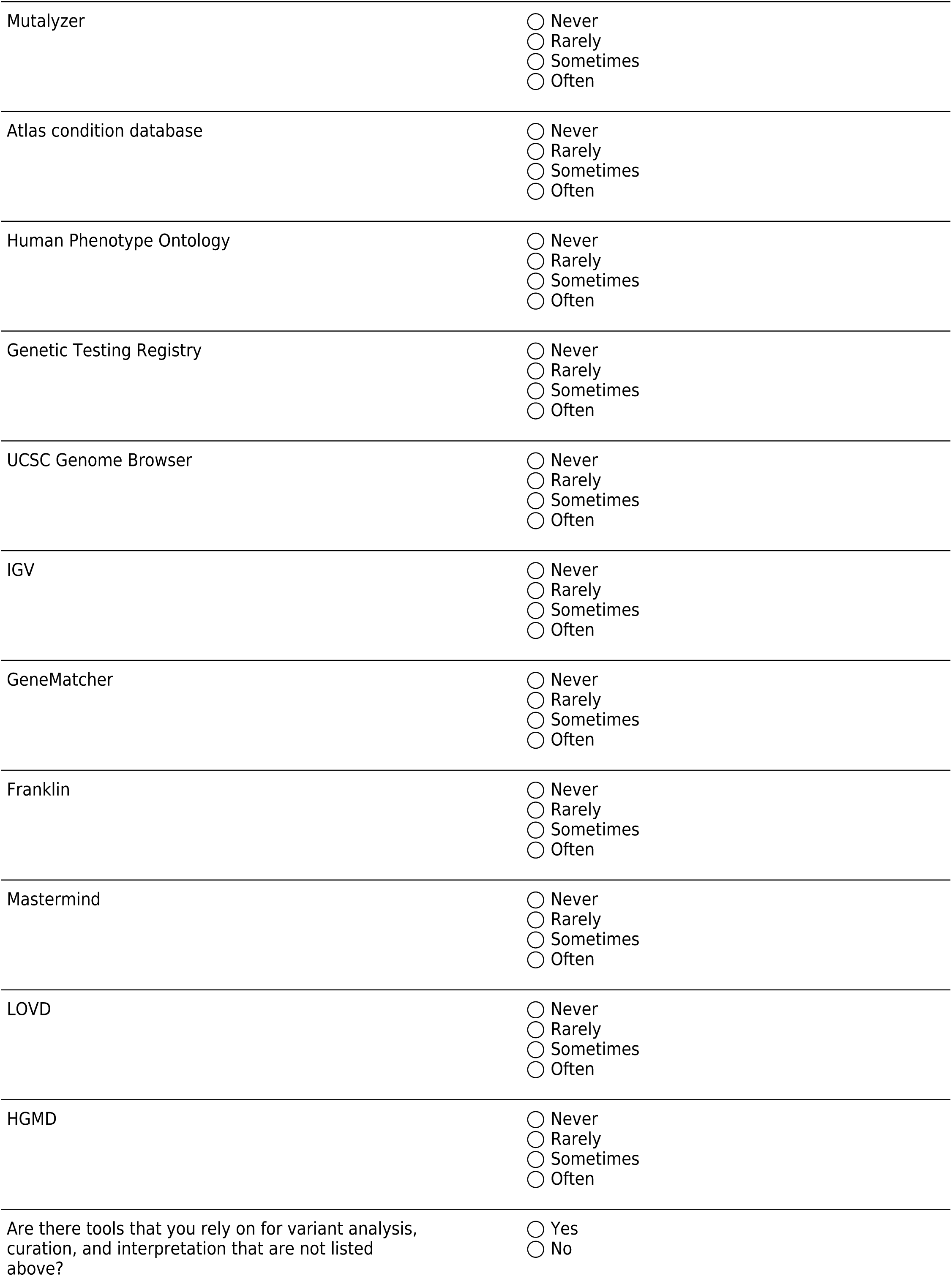
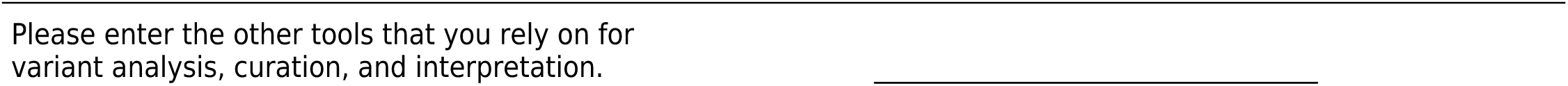

## REFERENCES

1. Kingsmore SF, Cole FS. The Role of Genome Sequencing in Neonatal Intensive Care Units. Annu Rev Genomics Hum Genet. 2022;23:427–48.

2. Malinowski J, Miller DT, Demmer L, Gannon J, Pereira EM, Schroeder MC, et al. Systematic evidence-based review: outcomes from exome and genome sequencing for pediatric patients with congenital anomalies or intellectual disability. Genet Med. 2020;22(6):986–1004.

3. Group NIS, Krantz ID, Medne L, Weatherly JM, Wild KT, Biswas S, et al. Effect of Whole-Genome Sequencing on the Clinical Management of Acutely Ill Infants With Suspected Genetic Disease: A Randomized Clinical Trial. JAMA Pediatr. 2021;175(12):1218–26.

4. Bowling KM, Thompson ML, Finnila CR, Hiatt SM, Latner DR, Amaral MD, et al. Genome sequencing as a first-line diagnostic test for hospitalized infants. Genet Med. 2022;24(4):851–61.

5. Austin-Tse CA, Jobanputra V, Perry DL, Bick D, Taft RJ, Venner E, et al. Best practices for the interpretation and reporting of clinical whole genome sequencing. NPJ Genom Med. 2022;7(1):27.

6. Richards S, Aziz N, Bale S, Bick D, Das S, Gastier-Foster J, et al. Standards and guidelines for the interpretation of sequence variants: a joint consensus recommendation of the American College of Medical Genetics and Genomics and the Association for Molecular Pathology. Genet Med. 2015;17(5):405–24.

7. Li MM, Datto M, Duncavage EJ, Kulkarni S, Lindeman NI, Roy S, et al. Standards and Guidelines for the Interpretation and Reporting of Sequence Variants in Cancer: A Joint Consensus Recommendation of the Association for Molecular Pathology, American Society of Clinical Oncology, and College of American Pathologists. J Mol Diagn. 2017;19(1):4–23.

8. Jenkins BD, Fischer CG, Polito CA, Maiese DR, Keehn AS, Lyon M, et al. The 2019 US medical genetics workforce: a focus on clinical genetics. Genet Med. 2021;23(8):1458–64.

9. Pirzadeh SM, McCarthy Veach P, Bartels DM, Kao J, Leroy BS. A national survey of genetic counselors’ personal values. J Genet Couns. 2007;16(6):763–73.

10. Maiese DR, Lyon M, Reddi HV, Blitzer MG, Bodurtha JN, Muenke M. The 2019 medical genetics workforce: A focus on laboratory geneticists. Genet Med. 2023;25(6):100834.

11. National Society of Genetic Counselors. 2024 Professional Status Survey. www.nsgc.org/Policy-Research-and-Publications/Professional-Status-Survey.

12. Rehm HL, Berg JS, Brooks LD, Bustamante CD, Evans JP, Landrum MJ, et al. ClinGen--the Clinical Genome Resource. N Engl J Med. 2015;372(23):2235–42.

13. Rivera-Munoz EA, Milko LV, Harrison SM, Azzariti DR, Kurtz CL, Lee K, et al. ClinGen Variant Curation Expert Panel experiences and standardized processes for disease and gene-level specification of the ACMG/AMP guidelines for sequence variant interpretation. Hum Mutat. 2018;39(11):1614–22.

14. ClinGen. Variant Pathogenicity Training Materials. www.clinicalgenome.org/curation-activities/variant-pathogenicity/training-materials.

